# A Generative Model For Evaluating Missing Data Methods in Large Epidemiological Cohorts

**DOI:** 10.1101/2024.04.23.24306030

**Authors:** Lav Radosavljević, Stephen M. Smith, Thomas E. Nichols

## Abstract

**Background:** The potential value of large scale datasets is constrained by the ubiquitous problem of missing data, arising in either a structured or unstructured fashion. When imputation methods are proposed for large scale data, one limitation is the simplicity of existing evaluation methods. Specifically, most evaluations create synthetic data with only a simple, unstructured missing data mechanism which does not resemble the missing data patterns found in real data. For example, in the UK Biobank missing data tends to appear in blocks, because non-participation in one of the sub-studies leads to missingness for all sub-study variables.

**Methods:** We propose a method for generating mixed type missing data mimicking key properties of a given real large scale epidemiological data set with both structured and unstructured missingness while accounting for informative missingness. The process involves identifying sub-studies using hierarchical clustering of missingness patterns and modelling the dependence of inter-variable correlation and co-missingness patterns.

**Results:** On the UK Biobank brain imaging cohort, we identify several large blocks of missing data. We demonstrate the use of our method for evaluating several imputation methods, showing modest accuracy of imputation overall, with iterative imputation having the best performance. We compare our evaluations based on synthetic data to an exemplar study which includes variable selection on a single real imputed dataset, finding only small differences between the imputation methods though with iterative imputation leading to the most informative selection of variables.

**Conclusions:** We have created a framework for simulating large scale data with that captures the complexities of the inter-variable dependence as well as structured and unstructured informative missingness. Evaluations using this framework highlight the immense challenge of data imputation in this setting and the need for improved missing data methods.

## 1 Background

Missing data is common in epidemiological and health data research and presents formidable challenges for many analytical approaches. The causes of missing data vary, from being inherent to the study design, to elective non-participation, or simply faults in measurement. Much work has therefore been devoted to evaluating the performance of methods for handling missing data. The most common approaches to comparing imputation methods include simulating data and inducing missingness using *a priori* chosen mechanisms^1,2^. Alternatively, artificial missingness is induced on real, complete data^3–5^ or real missingness patterns are imposed on simulated data^6^. Simulation studies of this kind usually rely on quite restrictive assumptions that might not be reflective of large scale epidemiological cohorts such as UK Biobank (UKB). For example, while some studies induce missingness in an unstructured manner^1,4^, in UKB missing data caused by non-participation in a sub-study/questionnaire comes in “blocks”. Specifically, if a subset of participants do not participate in an extension of the core study, then all of these subjects will have missing entries for the variables of this extension, and (when the rows and columns of the subjects-by-variables matrix are suitably reordered) this will form a solid block of missing data. Since UKB and similar datasets consist of many different sub-studies and questionnaires, this is a crucial feature to consider when evaluating the performance of imputation methods.

While there has been work done on the evaluation of existing methods on data with structured missingness and the development of new methods for handling such data, it has been common to use a non data-driven method for inducing structured missingness^2,7,8^ or use real missingness patterns imposed on simulated data^6^. Assuming that the structured missingness is Missing Completely at Random (MCAR) is especially problematic in the case where it is created by non-participation, since we know that participants often are disproportionately healthy^9^, meaning that the data is not likely to be MCAR. Our aim is therefore to define a method of generating synthetic data which has the same properties as a given data set. We want the pattern to satisfy the following three criteria:

1. There is structured missingness^10^, blocks of missingness caused by non-participation in sub-studies, as well as unstructured missingness that is not in blocks and is attributable to any other cause.
2. Missingness is informative in the sense of MAR (Missing at Random), where there is a relationship between missingness in a given variable and the observed elements of other variables.
3. There is an association between inter-variable correlation and inter-variable missingness similarity, typically where tightly correlated variables are more likely to be jointly missing.

With our framework for simulating such synthetic data, we evaluate the performance of several imputation methods. We are motivated by associations between the brain imaging variables with health, demographic, behavioural and lifestyle variables in UK Biobank. Thus we consider the subset of ≈ 40 000 subjects with imaging derived phenotype (IDP) data, and a collection of ≈ 20 000 non-Imaging Derived Phenotypes (nIDPs) variables; these nIDP variables are a mixture of continuous and binary variables (some of the binary variables are 1-hot encoding of categorical variables).

## 2 Methods

### 2.1 Terminology

Let *n* and *d* be the number of subjects and variables respectively, **X** be our *n* × *d* dataset and **M** be the *n* × *d* missingness matrix where *M*_*ij*_ = 1 if variable *j* is missing for subject *i* and *M*_*ij*_ = 0 if it is not missing. The following definitions and notation are central to our work:

1. **Variable-wise missingness pattern**. For any variable *j* = 1, 2, …, *d*, the *variable-wise missingness pattern* for variable *j* is 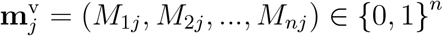.
2. **Subject-wise missingness pattern**. For any subject *i* = 1, 2, …, *n*, the *subject-wise missingness pattern* for subject *i* is 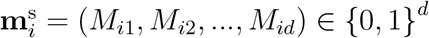.
3. **Variable-wise missingness distance**. For any two variables *j* and *j*^′^, the *variable-wise missingness distance* between them is the proportion of discordant missingness indicators

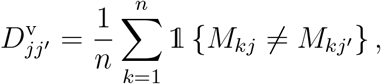

where 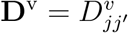 is the *d* × *d* variable-wise missingness distance matrix.
4. **Subject-wise missingness distance**. For any two subjects *i* and *i*^′^ the *subject-wise missingness distance* between them is likewise

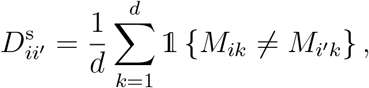

where **D**^s^ is the *n* × *n* subject-wise missingness distance matrix.
5. **Structured missingness**. We call missingness that is caused by non-participation in a sub-study/questionnaire *structured missingness*, resulting in a subset of subjects having missing data for a set of variables. This is also sometimes called block-wise missingness, as when subjects and variables are suitably reordered, this will result in solid blocks of missing data in the data matrix.
6. **Unstructured missingness**. We call missingness that is not caused by non-participation in a study/questionnaire *unstructured missingness*. This type of missingness will not induce any sort of blocks of missingness.

We now define the stochastic mechanisms that can give rise to missing data. Let **x** be a *d*-dimensional random vector drawn from the same distribution as the data in our data set and **m** be its corresponding subject-wise missingness pattern. Let further **x**_obs(**m**)_ and **x**_miss(**m**)_ be the observed and unobserved parts of the random vector **x** respectively. We follow the terms in Rubin (1976) for different types of missingness:

- *Missing Completely at Random* (MCAR)

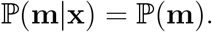 This means that the missingness mask is completely independent from underlying data.
- *Missing at Random* (MAR)

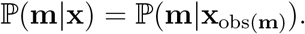 This means that there exists some dependence between the missingness mask and the underlying data, but that this relationship can be described using only observed data, i.e., the relationship between **m** and **x** is determined exclusively by the observed part **x**_obs(**m**)_. For example, this can mean that there exists a group of variables with no missingness which determine the missingness mask **m**.
- *Missing Not at Random* (MNAR)

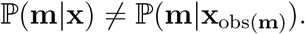 For this type of missingness, the relationship between the data and the missingness requires knowledge of underlying data. This is the most difficult setting to handle, assuming no prior knowledge of the mechanism by which missingness is induced, since it has been shown that for any MNAR model explaining missing data in a given data set, there exists an MAR model with equal evidence ^11^. In other words, there can be no theoretical guarantees of correctness for MNAR models explaining missing data barring direct knowledge of the missingness mechanism.

Characterising types of missingness is crucial to our work since many methods of handling missing data, most notably Multivariate Imputation by Chained Equations (MICE), have theoretical guarantees under MCAR and MAR^12^, while MNAR requires additional assumptions^13^.

### 2.2 Parameters of the Generative Model

We assume that our data consists of *C* different sub-studies, where study *c* = 0 is assumed to be a baseline study with no missingness while studies *c* = 1, …, *C* − 1 are follow up substudies with both structured and unstructured missingness. The following parameters define our generative model:

- {*ρ*}_*c,c′*_, the distribution of between-variable correlations for all pairs of clusters *c, c*^′^. We assume a mixed data generative model^14^, where data arise from a multivariate
- normal distribution with zero means and unit variances: for continuous variables these values are directly observed, while for binary variables the normal variate is latent and the data is obtained by thresholding continuous variables to 0*/*1. Therefore, {*ρ*}_*c,c′*_ represents the correlation distribution of the underlying data prior to thresholding.
- *π*_*c*_ the rate of structured missingness for cluster *c*.
- (*α*_*c*_, *β*_*c*_), parameters governing the rate of unstructured missingness for each variable. We assume that the rate of unstructured missingness 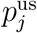 is drawn from Beta(*α*_*c*_, *β*_*c*_) if feature *j* belongs to sub-study *c*.
- Σ_core_. We assume that *d*_core_ variables from the baseline study *c* = 0 determine all structured missingness through a logistic model. Σ_core_ is the correlation matrix of these core variables. The core variables are assumed to all be continuous.
- AUC_*c*_, the Area Under the Curve (AUC) score of the logistic model determining structured missingness for sub-study *c*.

### 2.3 Estimating Parameters

We estimate the parameters of the model using the following procedure, also detailed in the flowchart in Figure 1.

**Figure 1:**
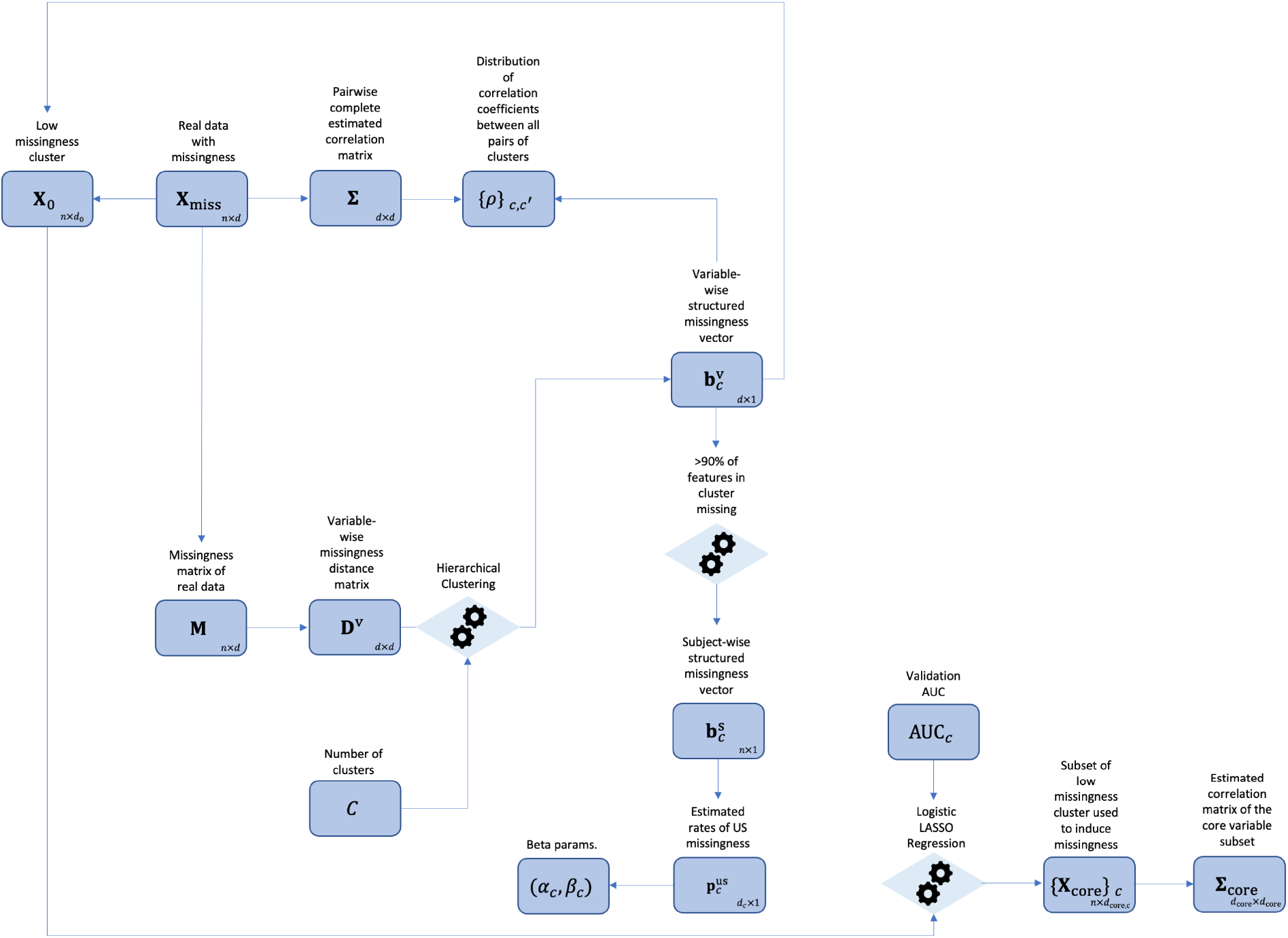
Flow chart of the data analysis pipeline.

1. Our *C* sub-studies are identified using hierarchical agglomerative complete linkage clustering^15^.
2. The densities {*ρ*}_*c,c′*_ are estimated using a histogram for each pair of clusters *c, c*^′^.
3. We define a subject *i* to be structurally missing for a sub-study *c* if at least 90% of the variables from *c* are missing for subject *i*. This will give us the vectors 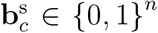 where 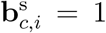 if subject *i* is structurally missing for sub-study *c*. This result also directly gives us *π*_*c*_, i.e., the probability of a subject having structured missingness for cluster *c*.
4. Having identified all structured missingness, we can estimate (*α*_*c*_, *β*_*c*_) using the method of moments on the remaining, unstructured missingness.
5. We use LASSO Logistic Regression (LASSO-LR)^16^ to simultaneously identify the core variables that determine structured missingness and AUC_*c*_, by fitting *C* − 1 penalised logistic regression models that use the baseline study data **X**_0_ as predictors and the subject-wise structured missingness vectors 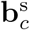 as outcomes. Specifically, AUC_*c*_ is estimated using 5-fold cross validation. Note that the core variables are cluster specific and may or may not overlap for different clusters. We then estimate Σ_core_, the correlation matrix of the core variables for all substudies. The penalty term *λ*_*c*_ for each LASSO models can be chosen in multiple appropriate ways (see subsection 2.4.1).

### 2.4 Generating Synthetic Data

The data is generated using a step-wise procedure as seen in Figure 2. Since all continuous variables have unit variance correlation matrices and covariance matrices of continuous data are the same.

**Figure 2:**
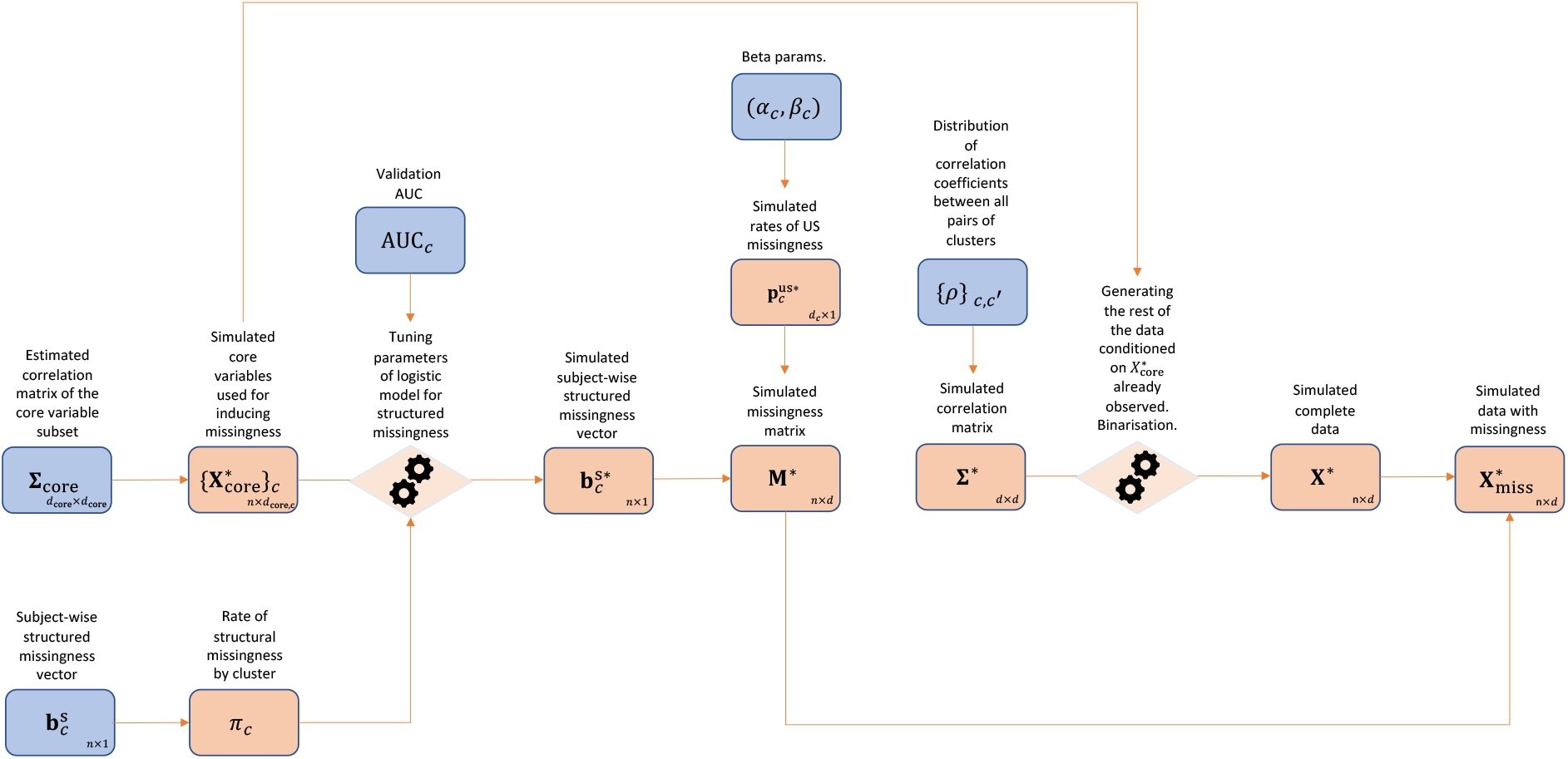
Flow chart of the synthetic data generation pipeline.

1. Using Σ_core_, we simulate the cluster specific core variables 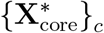 for clusters *c* = 1, …, *C* − 1, by drawing the full core variable data matrix 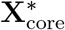 from 𝒩(**0**, Σ_core_).
2. Using a binary search procedure, we determine intercepts and coefficients of *C* − 1 logistic models determining structured missingness, with 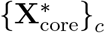 as covariates, such that the model’s AUC score and rate of positive cases will match AUC_*c*_ and *π*_*c*_. All the coefficients of the logistic models are assumed to be equal. Using these models, we generate synthetic subject-wise structured missingness vectors 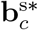.
3. We generate the rates of unstructured missingness 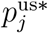 by drawing them independently from Beta(*α*_*c*_, *β*_*c*_) where variable *j* is in sub-study *c*. Unstructured missingness is assumed to be MCAR and is induced for each subject *i* and variable *j* with probability 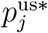. By combining the generated structured and unstructured missingness, we obtain an *n* × *d* synthetic missingness indicator matrix **M**^*^.
4. We simulate the full *d* × *d* correlation matrix Σ^*^ by drawing its entries from {*ρ*}_*c,c′*_ and, if necessary, projecting it to the nearest positive definite correlation matrix using Higham’s algorithm ^17^.
5. Having the complete correlation matrix, we generate the rest of the data **X**^*^, conditioned on the already simulated core variable data 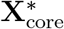. To allow for binary variables, we threshold a subset of variables to become binary, corresponding to the same number of binary variables in each cluster.
6. Finally the synthetic missingness mask **M**^*^ is imposed upon **X**^*^ to obtain the corresponding synthetic data set with missingness 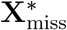.

This procedure generates synthetic datasets which satisfy the key criteria outlined in the introduction. Crucially, we have access to the true mean vector and covariance matrix, as well as the underlying data obscured by missingness.

#### 2.4.1 Calibrating the Predictability of Missingness

The choice of penalty term *λ*_*c*_ for each LASSO-LR model that predicts structured missingness can most easily be made by selecting the value of *λ*_*c*_ which minimises validation loss. This is, however, not always the option which is most faithful to the assumptions of our generative model. Since our generative model assumes that all core variables have equal importance in predicting structured missingness, we want to choose a value of *λ*_*c*_ which will minimise the number of core variables of low predictive importance, while having a validation loss that is close to that of the optimal value. This is an inevitably arbitrary feature of our generative model and we choose to select a reasonable value of *λ*_*c*_ through trial and error.

### 2.5 Simulation Study

In order to demonstrate the use of our generative model, we will conduct a simulation study on synthetic data mimicking the UK Biobank Brain nIDPs to evaluate the performance of three commonly used imputation methods on this data set. Our study will test the accuracy of imputation as measured by Mean Squared Error (MSE) for continuous variables and Balanced Accuracy (BA) for binary variables over *B* = 20 synthetically generated datasets. Additionally, to illustrate the difficulty of imputing data with structured missingness, we will perform the same simulation study on copies of the synthetic datasets where missingness has been induced in an MCAR and completely unstructured manner. The data in each sub-study is set to be missing using independent Bernoulli variables with probability equal to the total rate of missingness for the sub-study.

The first imputation method is mean imputation, which will serve as our benchmark method. The second is the matrix completion method SoftImpute^18^, which assumes that there exists a low rank approximation of the data set. This method has a tuning parameter, the value of the low rank, which we vary as 5%, 15% and 30% of the full matrix rank. Both the mean imputation and SoftImpute methods will be binarised to impute binary variables using 0.5 as the threshold, so imputed values greater than or equal to 0.5 will be transformed to 1, while the rest will be transformed to 0. The last method is called iterative imputation^19^ or ICE, Iterative Imputation by Chained Equations ^3^, which uses the same iterative procedure as MICE, but does not include randomness in the imputed values and only creates a single imputed data set. By testing the accuracy of iterative imputation we are effectively evaluating the accuracy of the “signal” component of the MICE imputation method. Additionally, MICE is impractical to use in this high dimensional setting with respect to memory use and computational time since it requires a large number of multiply imputed data sets. We chose to impute continuous values using Bayesian Ridge Regression^19^ and binary values using Logistic Regression with a Ridge penalty. In high dimensional setting, iterative imputation requires us to choose a subset of *k << d* variables that will be used to impute each variable *j*. These variables are normally set to be the *k* variables with highest absolute correlation with *j* ^19–21^, or select the *k* variables with the most favorable missingness patterns^20^; all else being equal, we favour variables for imputing *j* which are observed the most often in rows where *j* is missing and therefore select variables using the rows of the matrix

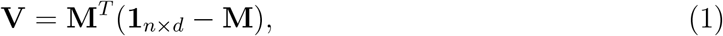

where *V*_*jj′*_ is the number of times *j*^′^ is observed when *j* is missing.

We propose a third selection method which utilises correlation and missingness jointly, while being applicable to mixed data. It calculates a score *S*_*jj′*_ which is proportional to the maximum expected reduction imputation error (MSE for continuous and misclassification rate for binary variables) under the assumption of MCAR and under the generative model described in^14^ for joint continuous and binary data, i.e., an underlying multivariate normal distribution with thresholding for binary variables.

- *j* **and** *j*^′^ **are both continuous**

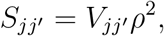

where *ρ* is the Pearson correlation between variables *j* and *j*^′^.
- *j* **is continuous and** *j*^′^ **is binary**

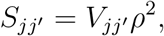

where *ρ* is the Pearson correlation between variables *j* and *j*^′^.
- *j* **is binary and** *j*^′^ **is continuous**

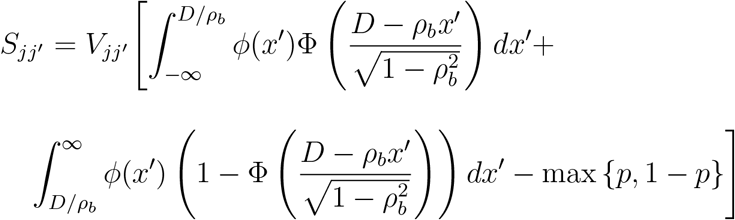

if *ρ*_*b*_ *>* 0 and

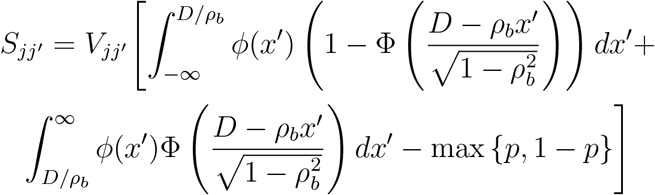

if *ρ*_*b*_ *<* 0, where *ϕ* and Φ are the the probability density function and cumulative distribution function of the standard-normal distribution, *ρ* is the Pearson correlation between variables *j* and *j*^′^, *p* is the rate of positive cases for variable *j, D* = Φ^−1^(*p*) and

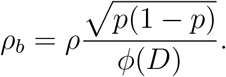

*D* is the threshold of the underlying standard-normal variable that determines the binary value of *j* and *ρ*_*b*_ is the correlation between this underlying variable and *j*^′^. The reduction in misclassification loss can be calculated directly using these quantities by assuming that we predict 0*/*1 depending on whether the median of the latent variable conditioned on the value of *j*^′^ is greater than *D* or not.
- *j* **and** *j*^′^ **are both binary**

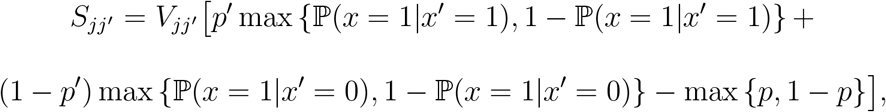

where *p* and *p*^′^ are the rates of positive cases for *p* and *p*^′^ respectively. Here, the reduction in misclassification loss is calculated directly from the 2 × 2 contingency table of *j* and *j*^′^, since we know the most likely outcome of variable *j* given the value of *j*^′^. This contingency table is calculated using *p, p*^′^ and *ρ*.

A formal proof of these results can be found in the supplementary material. We will vary the tuning parameter *k* to be 10, 50 and 150.

### 2.6 Illustrative Example: Variable Selection for Predicting Total Grey Matter Volume

In order to demonstrate the validity of the conclusions drawn from our simulation study, we apply our imputation methods to an analytical task on real data and see if there is agreement between the results of the analysis and the conclusions drawn from the simulation study. We chose the task of selecting 15 nIDPs for an Ordinary Least Squares (OLS) model predicting log-transformed normalised total grey matter volume. The total pool to select from is ≈ 15 000 nIDPs (nIDPs with 0 variance or missingness above 40% were excluded). Imputation is used here as a pre-processing step and LASSO-LR is used for variable selection. The outcome, i.e., the 15 variables that are selected will vary depending on the imputation method. We compare four different approaches: using only complete varibles, mean imputation, SoftImpute and iterative imputation. The tuning parameters for SoftImpute and iterative imputation are chosen based on their performance in the simulation study. The four approaches are evaluated by the relevance of the 15 selected variables, as measured by the pooled *R*^2^ estimates of each OLS model. To ensure fair assessment of the *R*^2^ scores irrespective of missingness in the selected variables, we use the mice package in R^13^ to create *m* = 100 multiply imputed data sets of all selected variables and pool the *R*^2^ scores and their standard error estimates for each OLS model according to “Rubin’s rules” ^22^. We use the following estimator of this standard error ^23^:

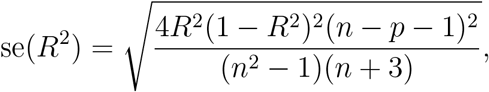

where *n* is the number of observations and *p* the number of variables.

We also ensure that the baseline variables age squared, sex and Townsend deprivation index are included in the OLS model as potential confounding variables.

## 3 Results

### 3.1 Analysis Pipeline

As seen in subsection 2.3, we need to select a value for the number of substudies/clusters *C* as a parameter of our analysis pipeline. This choice is jointly driven by the data itself as well as the need to select a small number of clusters/substudies to allow us to clearly illustrate our methodology. Figure 3 shows the dendrogram of the 100 last agglomerations in the hierarchical clustering of nIDPs by variable-wise missingness pattern. It can be seen from this dendrogram that *C* = 4 will give us clusters which have a high between-cluster missingness distance relative to within-cluster distance. The choice of *C* = 4 clusters also gives us reasonably sized clusters for our analysis, as seen in Table 1.

**Table 1:** Table of cluster sizes.

| Cluster | # of Binary Variables | # of Continuous Variables | Total # of Variables |
| --- | --- | --- | --- |
| $c = 0$ | 13458 | 1227 | 14685 |
| $c = 1$ | 371 | 6434 | 6805 |
| $c = 2$ | 131 | 2012 | 2143 |
| $c = 3$ | 3 | 235 | 238 |

**Figure 3:**
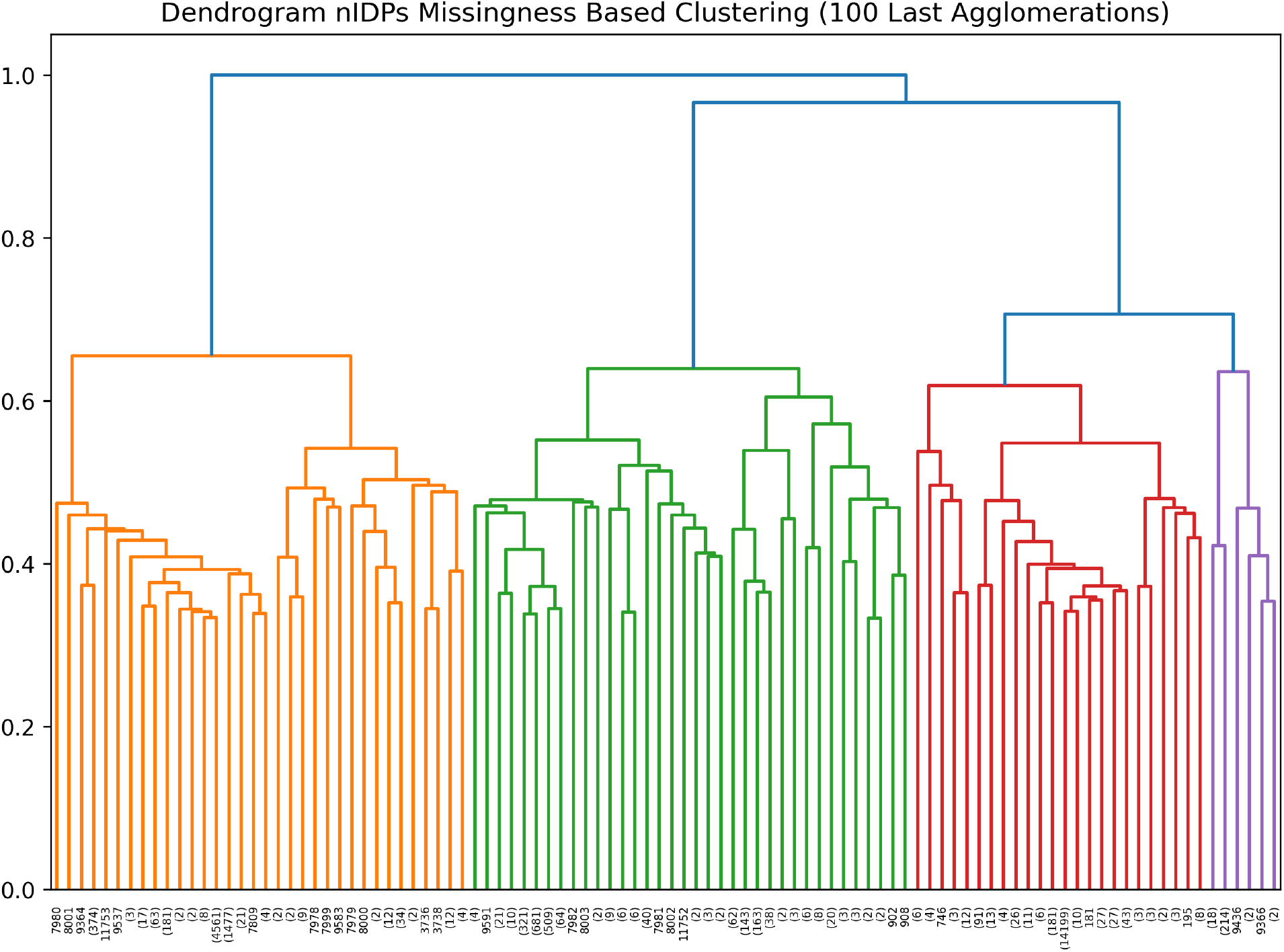
Dendrogram of the 100 last agglomerations in the hierarchical clustering of nIDPs by variable-wise missingness pattern, where distance between merged clusters (*y*-axis) is the maximal variable-wise missingness distance between agglomerated clusters. We determine that *C* = 4 clusters/substudies is an appropriate choice for illustrating the workings of our method since it gives us reasonably sized clusters with a high between-cluster distance relative to within-cluster distance.

The share of each nIDP type by cluster is shown in Figure 4. Cluster *c* = 0 contains almost exclusively health and medical related nIDPs, cluster *c* = 2 contains mostly lifestyle and environment related variables, cluster *c* = 3 almost exclusively contains cognitive phenotype variables and cluster *c* = 1 contains a mix of the remaining types of variables. This results shows that nIDPs of the same type tend to have similar variable-wise missingness patterns.

Figure 5 plots the histograms of the proportions of variable-wise missing data, i.e., fraction of subjects missing for each variable in a cluster. As we can see, the cluster *c* = 0, the cluster that contains mostly health- and medical related variables, has almost no missingness. This is an expected result, as health records usually either contain too much missingness to be included in the first place or they have very little missingness as absence of data indicates abscence of recorded disease or diagnosis. Clusters *c* = 2, 3 have intermediate rates of missingness with cluster *c* = 3 having lower rates of missingness as well as a lower variability of rates of missingness, while *c* = 1 has very high rates of missingness. For this reason, we will exclude cluster *c* = 1 from our generative model, as its variables have too high rates of missingness to be interesting to use for imputation.

**Figure 4:**
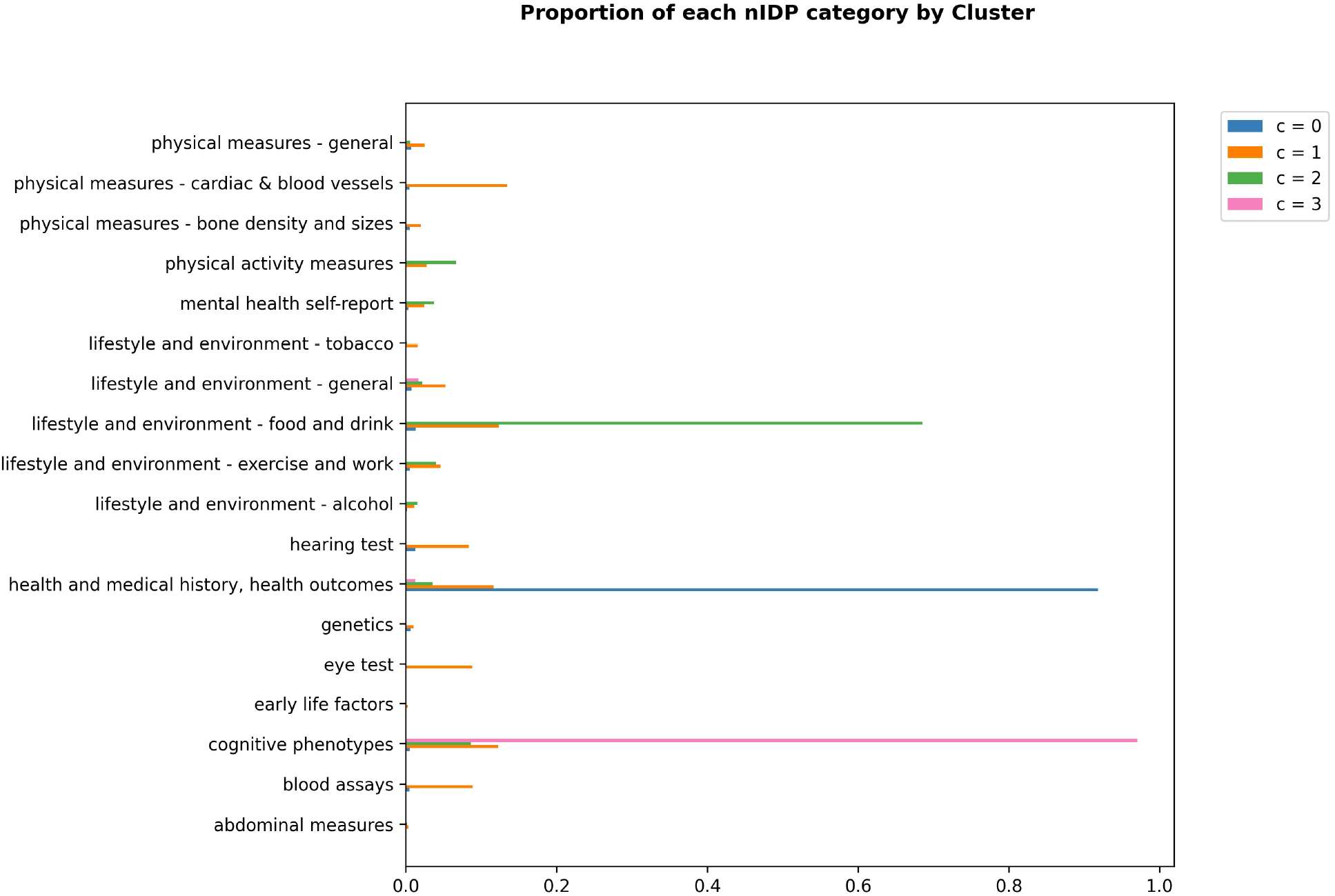
Bar plots detailing the proportion of nIDP variable types in each cluster. Cluster *c* = 0 contains almost exclusively health and medical related nIDPs, cluster *c* = 2 contains mostly lifestyle and environment related variables, cluster *c* = 3 almost exclusively contains cognitive phenotype variables and cluster *c* = 1 contains a mix of the remaining types of variables. This shows that nIDPs of the same type tend to have similar variable-wise missingness patterns.

**Figure 5:**
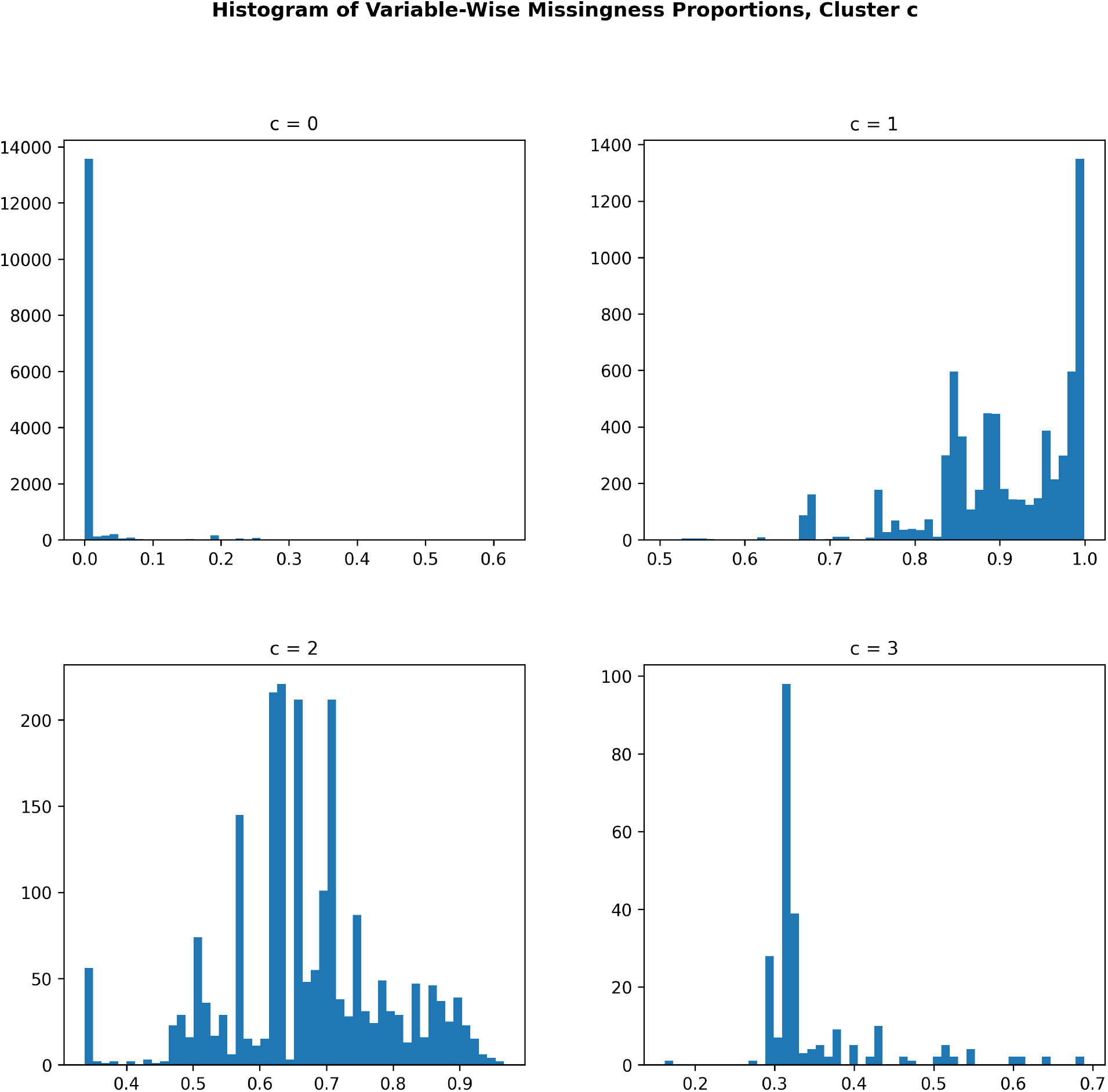
Histograms of the proportions of missing data for variables in each cluster. Each entry in the histogram for cluster *c* is the proportion of missing data for a single variable belonging to cluster *c*. Cluster *c* = 0, i.e., the cluster that contains mostly health- and medical related variables, has almost no missingness. Clusters *c* = 2, 3 have intermediate rates of missingness with cluster *c* = 3 having lower rates of missingness as well as a lower variability in the same, while *c* = 1 has very high rates of missingness.

Figure 6 displays the subject-wise missingness histograms, the proportion of cluster-*c* variables missing for a given subject. The red line in each plot signifies the 90% threshold for structured missingness, meaning that subjects for which 90% or more of the features assigned to cluster *c* are missing are considered to have structured missingness for the variables in cluster *c*. We can see that cluster *c* = 3 has a much clearer separation between structured and unstructured missingness, whereas it is less clear for clusters *c* = 1, 2, likely due to higher rates of unstructured missingness as well as our approximation of *C* = 4 leading to different clusters being grouped together.

**Figure 6:**
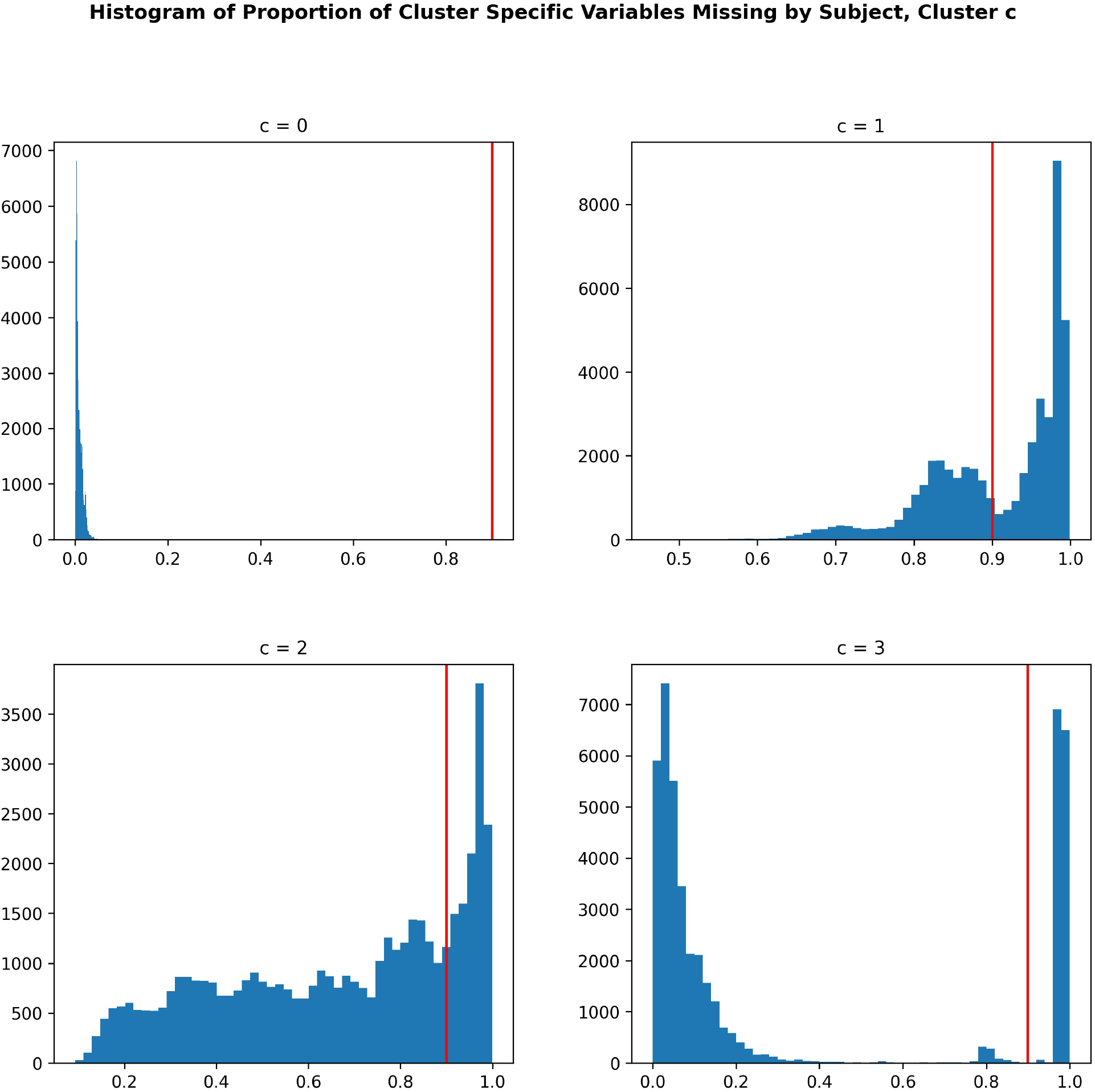
Histograms detailing the proportion of variables assigned to cluster *c* that are missing, by subject. The red line in each plot signifies the 90% threshold for structured missingness, meaning that subjects for which 90% or more of the features assigned to cluster *c* are missing are considered to have structured missingness for the variables in cluster *c*. We can see that the cluster *c* = 3 has a much clearer separation between structured and unstructured missingness, whereas it is less clear for clusters *c* = 1, 2, likely due to higher rates of unstructured missingness as well as our approximation of *C* = 4 leading to different

As discussed in subsection 2.4.1, the penalty terms *λ*_*c*_ need to be carefully chosen to not violate the assumptions of our generative model. Manual tuning arrived at a value of *λ*_*c*_ = exp(6) for both clusters *c* = 2, 3 which minimised both the total number of core variables as well as the proportion of binary variables, while having validation AUC scores close to those of the optimal values, as shown in Tables 2 and 3.

**Table 2:** Table detailing the validation AUC for predicting structured missingness using variables from cluster *c* = 0 and number of binary and continuous variables selected using LASSO Logistic Regression (LASSO-LR) for optimal values of *λ*_*c*_.

| Cluster | AUC<sub>c</sub> | # of Binary Variables | # of Continuous Variables | $d_{\text{core},c}$ |
| --- | --- | --- | --- | --- |
| $c = 2$ | 0.72 | 48 | 575 | 623 |
| $c = 3$ | 0.90 | 271 | 948 | 1219 |

**Table 3:** Table detailing the validation AUC for predicting structured missingness using variables from cluster *c* = 0 and number of binary and continuous variables selected using LASSO logistic regression using *λ*_*c*_ = exp(6) for both clusters *c* = 2, 3.

| Cluster | AUC<sub>c</sub> | # of Binary Variables | # of Continuous Variables | $d_{\text{core},c}$ |
| --- | --- | --- | --- | --- |
| $c = 2$ | 0.71 | 5 | 62 | 67 |
| $c = 3$ | 0.86 | 5 | 78 | 83 |

The final results of the data analysis are summarised in Table 4. These results indicate that clusters *c* = 2, 3 have similar rates of structured missingness, while cluster *c* = 2 has a much higher rate of unstructured missingness, as indicated by the values of *α*_*c*_ and *β*_*c*_. It is also apparent that the variables in cluster *c* = 3 have a more informative type of structured missingness as we can see by the higher value of AUC_*c*_.

**Table 4:** Table of results summarising the data analysis step of the method. These results indicate that clusters *c* = 2, 3 have similar rates of structured missingness, while cluster *c* = 2 has a much higher rate of unstructured missingness, as indicated by the values of *α*_*c*_ and *β*_*c*_. It is also apparent that the variables in cluster *c* = 3 have a more informative type of structured missingness as we can see by the values of AUC_*c*_.

| Cluster | $\text{AUC}_c$ | $d_{\text{core},c}$ | $\pi_c$ | $\alpha_c$ | $\beta_c$ |
| --- | --- | --- | --- | --- | --- |
| $c = 2$ | 0.71 | 67 | 0.26 | 6.0 | 4.6 |
| $c = 3$ | 0.86 | 83 | 0.29 | 0.44 | 4.4 |

### 3.2 Simulation Study

Figure 7 plots the imputation accuracy by variable for datasets using the generative model as well as the unstructured equivalent described in sub-section 2.5. The red line in each violin plot is the median of the best performing method in that comparison, i.e., the lowest median MSE and the highest median BA. To interpret the results, it should be noted that the continuous variables of the generative model all have zero mean and unit variance. Iterative imputation that uses Pearson correlation or the mixed score as its criterion for variable selection is the best performing method overall. It is also notable that the SoftImpute performs poorly for binary data and notably worse than the iterative imputation methods for continuous data. When comparing the performance between generative model versus completely unstructured missing data, we can see that performance is better for the completely unstructured case, for both continuous and binary variables. This difference is particularly stark for cluster *c* = 3, where there is a lot of structured missingness and very little unstructured missingness, highlighting the difficulty of imputation in this setting. Finally, it should be noted that we are rarely able to explain more than 20% of variance in the missing values and that this could mean that the choice of imputation method will not greatly impact the outcome of many analytical tasks, as the modest accuracy of imputation may not be enough to greatly alter the final outcome.

**Figure 7:**
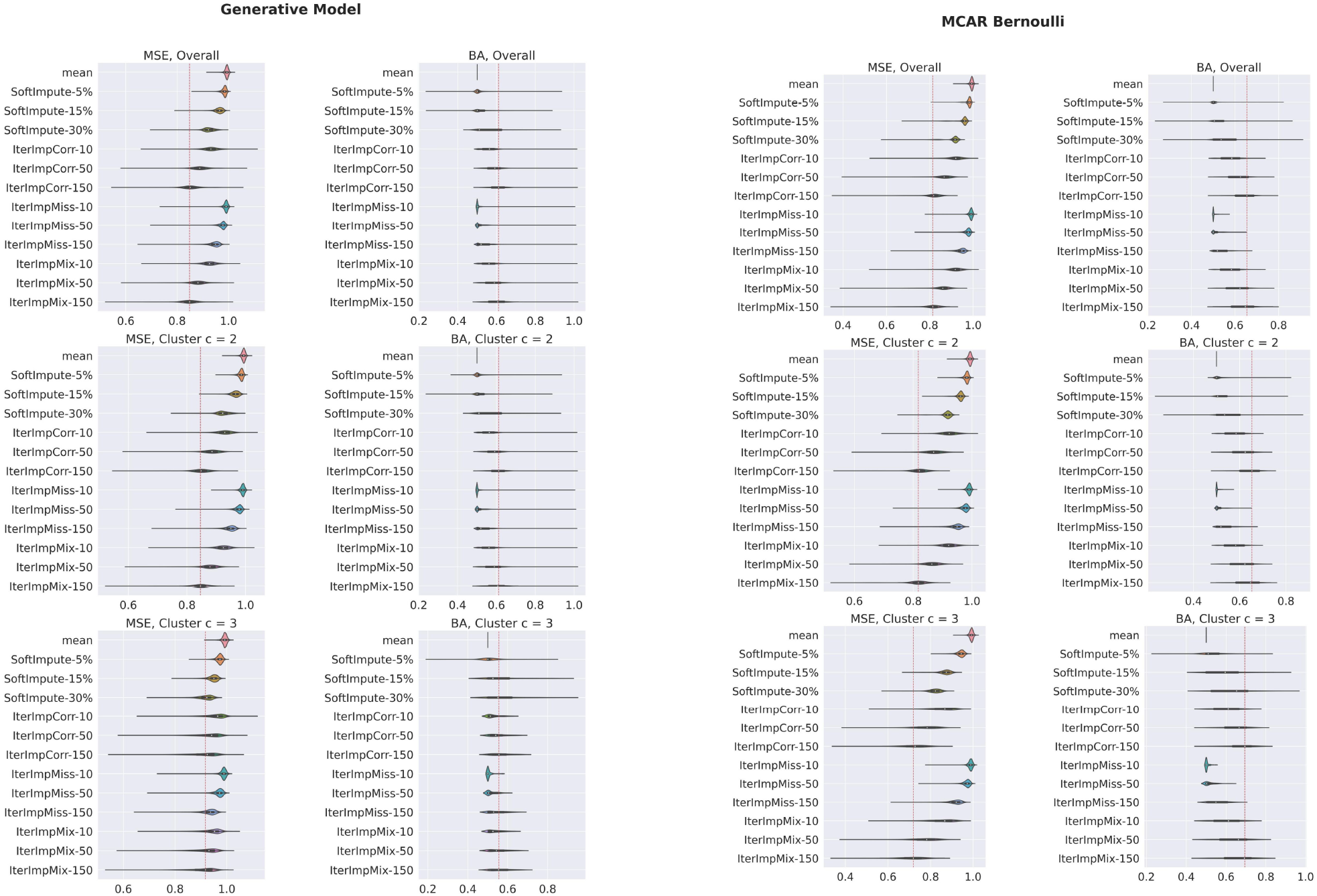
Violin plots of the imputation accuracy by variable. The red line in each violin plot is the median of the best performing method in that comparison, i.e., the lowest median MSE and the highest median BA. Since continuous data is standardised, the MSE scores correspond to 1 minus the variance explained by prediction. It is obvious that iterative imputation that uses Pearson correlation or the mixed score as its criterion for selecting *k* variables is the best performing method overall. It is also notable that SoftImpute performs poorly for binary data. When comparing the performance between generative model versus completely unstructured missing data, we can see that performance is better for the completely unstructured case, for both continuous and binary variables. This difference is particularly stark for cluster *c* = 3, where there is a lot of structured missingness and very little unstructured missingness, highlighting the difficulty of imputation in this setting.

### 3.3 Illustrative Example

Table 5 lists the variance explained for the OLS model using the selected variables along with the number of variables in the model that ended up being statistically significant. The results show a modest difference between the three imputation methods, with iterative imputation having the most statistically significant variables and the best *R*^2^ score. The results when using only complete variables are worse with a considerably lower *R*^2^ score as well as fewer variables ending up statistically significant. We also see that the complete variables method selected many more binary variables and this is because the complete variables are mostly health record data, i.e., data assigned to cluster *c* = 0, which is disproportionately binary as seen in Table 1. Meanwhile, the results for iterative imputation are the best, having the highest *R*^2^ score as well as the highest number of statistically significant variables. These results align well with our simulation study; a small difference in the final outcome of the analytical task for different methods caused due to the difficulty of imputing structurally missing data, but with iterative imputation clearly being the best alternative.

**Table 5:**
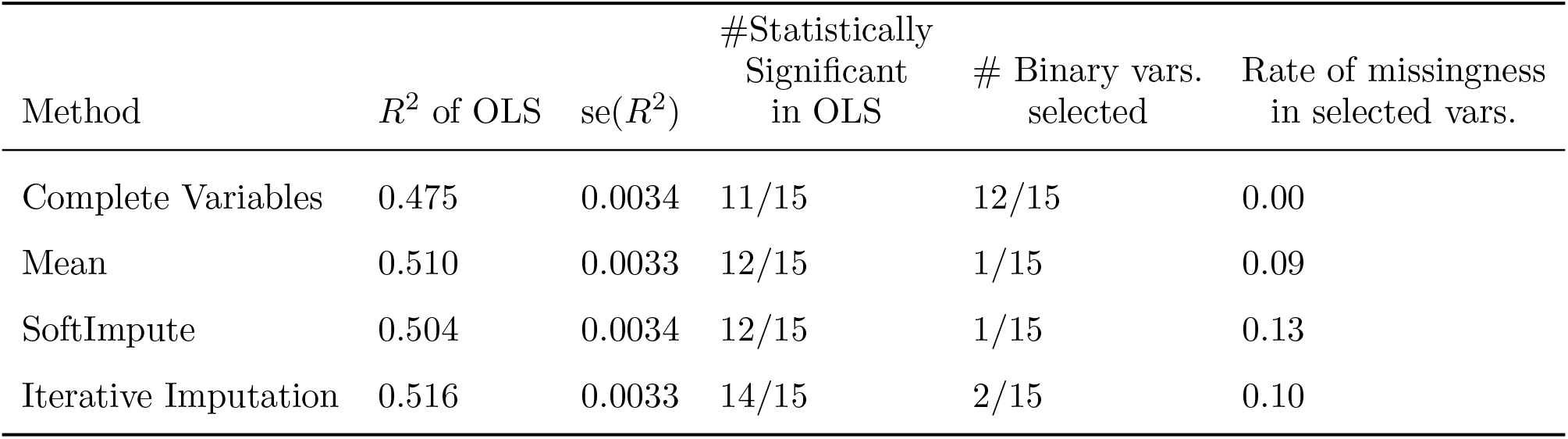
*R*^2^ scores for the OLS models using the selected variables along with the number of variables in the model that ended up being statistically significant. The results show a modest difference between the three imputation methods, with iterative imputation having the most statistically significant variables and the best *R*^2^ score. The results when using only complete variables are worse with a considerably lower *R*^2^ score as well as fewer variables ending up statistically significant.

## 4 Discussion

We have proposed a method for generating large-scale data with complex patterns of missing data that make imputation difficult. In particular, our data-driven simulation framework allows for highly informative missingness and joint missingness for variables that are strongly correlated. This ability to mimic the properties of large scale epidemiological datasets makes our method useful for gaining insight into the performance of handling missing data for different analytical tasks. There are, however, limitations to our model which are important to note and represent potential future work on this topic. Our model assumes multivariate normality for all continuous features, which limits the generalisability of any conclusions drawn from simulation studies of analytical methods that are sensitive to non-gaussianity or strong outliers. In such scenarios, it is possible that conclusions drawn using our generative model would unduly favour linear methods over more complicated black-box methods that would fare better on real, non-Gaussian data. This could be solved by parameterising the model differently, allowing for more flexibility on the underlying multivariate distribution, or by using non-parametric methods.

Another potential limitation of our generative model is that we assume that the correlation structure of the data closely follows the missingness structure. This is because we assume that for any pair of clusters *c, c*^′^, the correlation between pairs of variables in *c* and *c*^′^ are drawn independently from some distribution {*ρ*}_*c,c′*_, i.e., we assume that there is no further covariance structure within or between sub-studies. We have found this to be approximately true for the nIDPs that we have been working with, but this might not be the case for other datasets. This could be solved by modelling missingness and correlation structure jointly in a way which allows for further complexity inside sub-studies.

In this paper we chose to use *C* = 4 clusters of variables as an approximation of reality in order to be able to inspect the properties of these clusters separately. In all likelihood, the true number of substudies is higher, and more representative results could be obtained by choosing a higher figure. We deemed it necessary to use a lower number in order to demonstrate the inner workings of our method. When allowing *C* to be higher, we found *C* = 15 clusters with 100 or more variables present in the data set. These clusters all had a very clear separation between the structured and unstructured missingness, which bolsters our hypothesis that missingness in UKB data can be effectively modelled as we have suggested.

## 5 Conclusions

The results from the simulation study combined with the illustrative example show that there is room for improvement in the current missing data methodology to accommodate for this specific type of missing data, and that the final result of many analytical tasks on data from the UKB Brain Imaging cohort will vary little depending which commonly used imputation method is chosen. This is due to the difficulty of imputation in this setting. Even so, we have shown some advantage in using iterative imputation over matrix completion methods. While we do not propose new missing data methods here, our results highlight the need for developing methods that specifically account for structured missingness.

## Supporting information

Supplemental Proof

## Data Availability

One example dataset generated using our method can be found here:
https://www.kaggle.com/datasets/lrstats/example-dataset-generative-model
Further data can be made available upon reasonable request to authors.

## List of Abbreviations

UKB: UK Biobank
nIDP: non-Imaging Derived Phenotype
MCAR: Missing Completely at Random
MAR: Missing at Random
MNAR: Missing not at Random
AUC: Area Under the (Reciever Operating Characteristic) Curve
LASSO-LR: Least Absolute Shrinkage and Selection Operator Logistic Regression
MICE: Multivariate Imputation by Chained Equations
ICE: Imputation by Chained Equations
MSE: Mean Squared Error
BA: Balanced Accuracy
OLS: Ordinary Least Squares

## Declarations

### Ethics approval and consent to participate

The UK Biobank has received ethical approval from the North West Multi-Center Research Ethics Committee (11/NW/0382).

This research project received approval from the UKB under application number 8107.

This research project has adhered to the Declaration of Helsinki

### Consent for publication

Not applicable.

### Availability of data and materials

All code can be found in the following GitHub repository: https://github.com/lavrad99/Generative_Model_Missing_Data/

One example dataset generated using our method can be found here: https://www.kaggle.com/datasets/lrstats/example-dataset-generative-model

Further data can be made available upon reasonable request to authors.

### Authors’ contributions

LR performed data analysis, simulation and the exemplar study. All authors contributed ideas throughout the project. All authors contributed to and revised the final manuscript.

## Acknowledgements

The computational aspects of this research were supported by the Wellcome Trust Core Award Grant Number 203141/Z/16/Z and the NIHR Oxford BRC. The views expressed are those of the author(s) and not necessarily those of the NHS, the NIHR or the Department of Health

