## Supplemental Proof for "A Generative Model For Evaluating Missing Data Methods in Large Epidemiological Cohorts"

### Supplementary Proof for the paper “A Generative Model For Evaluating Missing Data Methods in Large Epidemiological Cohorts”

Lav Radosavljevic

August 2023

Here is a supplementary proof for the proposed variable selection method for iterative imputation. The results assume the joint continuous and binary model described by Demirtas and Doganay in 2012, meaning that we assume that the data is drawn from a underlying multivariate normal distribution, where binary data is obtained through thresholding. For the sake of simplicity, we will assume that the latent normal variables that have been thresholded have unit variance and zero mean.

Let $j$ and $j'$ be two variables in our data set $\mathbf{X}$ with missing data and let $(x, x')$ be a generic random draw of the pair $j$ and $j'$ where $x$ is missing and $x'$ is observed. We want to use the variable $j'$ to impute the values in $j$. It is assumed that we know the Pearson correlation $\rho$ between them, the missingness matrix $\mathbf{M}$ and the proportion of positive cases if either of them are binary. We also assume that the missingness falls under MCAR, so the missingness and the underlying values are completely independent. We want to know how much better than mean/mode imputation it can possibly be to use variable $j'$ to impute variable $j$, using square- or misclassification loss depending on the type of variable $j$. We want to preface by observing that the expected reduction in MSE/increase in accuracy will be proportional to $V_{jj'}$, the number of times variable $j$ is missing while $j'$ is observed, multiplied by the expected reduction in loss using the single generic pair $(x, x')$, because using $j'$ to impute $j$ is only useful in those instances where $j$ is missing and $j'$ is not. It is also clear that

$$\mathbf{V} = (\mathbf{1}_{n \times d} - \mathbf{M})^T \mathbf{M}.$$

##### $x$ and $x'$ are both continuous

If $x$ and $x'$ are both continuous normal variables with correlation $\rho$ and $\mathbb{E}[x] = \mu$ and $\text{Var}[x] = \sigma^2$, then it is obvious that

$$\mathbb{E}[(x - \mu)^2] - \mathbb{E}[(x - f_{\text{Bayes}}(x'))^2] = \sigma^2 - (1 - \rho^2)\sigma^2 = \rho^2\sigma^2,$$

because we know that the Bayes predictor under the square loss for $x$ is the conditional expectation of $x$ given $x'$ and $\mathrm{Var}[x|x'] = (1-\rho^2)\sigma^2$. Therefore, the decrease in MSE for using $j'$ to impute $j$ is proportional to $V_{jj'}\rho^2$.

##### $x$ is continuous and $x'$ is binary

Let $f_{\mathrm{Bayes}}(x') = \theta_0 + \theta_1 x'$ be the Bayes predictor under the square loss when $x'$ is a binary r.v. This form can be assumed without loss of generality (WLOG), since $f_{\mathrm{Bayes}}$ is only defined on the set $\{0, 1\}$. Also, let $p'$ be the proportion of positive cases for $j'$. We know that

$$\mathbb{E}[xx'] = \rho\sqrt{\mathrm{Var}[x]\mathrm{Var}[x']} + \mathbb{E}[x]\mathbb{E}[x'] = \rho\sigma\sqrt{p'(1-p')} + p'\mu.$$

We also know that $f_{\mathrm{Bayes}}$ minimises the expression

$$\begin{gathered}\mathbb{E}[(x - f_{\mathrm{Bayes}}(x'))^2] = \\ = \mathbb{E}[x^2] - 2\mathbb{E}[x f_{\mathrm{Bayes}}(x')] + \mathbb{E}[f_{\mathrm{Bayes}}(x')^2] = \\ \mu^2 + \sigma^2 - 2\mu\theta_0 - 2(\rho\sigma\sqrt{p'(1-p')} + p'\mu)\theta_1 + (\theta_0 + \theta_1 p')^2 + \theta_1^2 p'(1-p').\end{gathered}$$

We find the minimum with respect to $\theta_{0,1}$ by finding the point at which the gradient vector is 0. We have that:

$$\frac{\partial}{\partial\theta_0}\mathbb{E}[(x - f_{\mathrm{Bayes}}(x'))^2] = -2\mu + 2(\theta_0 + \theta_1 p') = 0 \Longleftrightarrow \theta_0 + \theta_1 p' = \mu.$$

And also:

$$\frac{\partial}{\partial\theta_1}\mathbb{E}[(x - f_{\mathrm{Bayes}}(x'))^2] = -2(\rho\sigma\sqrt{p'(1-p')} + p'\mu) + 2p'(\theta_0 + \theta_1 p') + 2p'(1-p')\theta_1 = 0$$

$$\Longleftrightarrow$$

$$-\rho\sigma\sqrt{\frac{1-p'}{p'}} - \mu + \theta_0 + \theta_1 p' + (1-p')\theta_1 = 0$$

$$\Longleftrightarrow$$

$$\theta_0 + \theta_1 = \mu + \rho\sigma\sqrt{\frac{1-p'}{p'}}.$$

This gives us the following system of equations:

$$\begin{cases}\theta_0 + p'\theta_1 = \mu \\ \theta_0 + \theta_1 = \mu + \rho\sigma\sqrt{\frac{1-p'}{p'}},\end{cases}$$

which evaluates to

$$\begin{cases} \theta_0 = \mu - \rho\sigma\sqrt{\frac{p'}{1-p'}} \\ \theta_1 = \frac{\rho\sigma}{\sqrt{p'(1-p')}}. \end{cases}$$

Using these values for $f_{\text{Bayes}}(x') = \theta_0 + \theta_1 x'$, we get that

$$\begin{gathered} \mathbb{E}[(x - f_{\text{Bayes}}(x'))^2] = \\ = \mu^2 + \sigma^2 - 2\mu\left(\mu - \rho\sigma\sqrt{\frac{p'}{1-p'}}\right) - 2(\rho\sigma\sqrt{p'(1-p')} + p'\mu)\left(\frac{\rho\sigma}{\sqrt{p'(1-p')}}\right) + (\mu)^2 \\ + \left(\frac{\rho\sigma}{\sqrt{p'(1-p')}}\right)^2 p'(1-p') = \\ = \mu^2 + \sigma^2 - 2\mu^2 + 2\mu\rho\sigma\sqrt{\frac{p'}{1-p'}} - 2\rho^2\sigma^2 - 2\mu\rho\sigma\sqrt{\frac{p'}{1-p'}} + \mu^2 + \rho^2\sigma^2 = \\ = (1-\rho^2)\sigma^2. \end{gathered}$$

This is the same result as in the previous case, which means that the decrease in MSE for using $j'$ to impute $j$ is proportional to $V_{jj'}\rho^2$.

##### $x$ is binary and $x'$ is continuous

Here we assume that $\tilde{x}$ is the underlying normal variable variable that is binarised using threshold $D \in \mathbb{R}$, i.e., $x = \mathbb{1}\{\tilde{x} > D\}$. $(\tilde{x}, x')$ is multivariate normal distributed. We can further assume WLOG that $\tilde{x}$ and $x'$ have zero mean, unit variance and correlation $\rho_b$. It is obvious that $f_{\text{Bayes}}(x') = \mathbb{1}\{\mathbb{E}[\tilde{x}|x'] > D\}$ is the Bayes predictor of $x$ using $x'$ under the misclassification loss, since $\tilde{x}|x'$ is normally distributed with expectation $\mathbb{E}[\tilde{x}|x']$ and the expected value and median of normally distributed variables are the same. We also know that $\mathbb{E}[\tilde{x}|x'] = \rho_b x'$, $\text{Var}[\tilde{x}|x'] = 1 - \rho_b^2$. This means that

$$\begin{cases} f_{\text{Bayes}}(x') = \mathbb{1}\{x' > D/\rho_b\} & \text{if } \rho_b > 0 \\ f_{\text{Bayes}}(x') = \mathbb{1}\{x' < D/\rho_b\} & \text{if } \rho_b < 0. \end{cases}$$

The expected decrease in misclassification loss by imputing $x$ using $x'$ is

$$\mathbb{E}[\mathbb{1}\{x = f_{\text{Bayes}}(x')\}] - \mathbb{E}[\mathbb{1}\{x = \arg\max(1-p, p)\}] = \mathbb{E}[\mathbb{1}\{x = f_{\text{Bayes}}(x')\}] - \max\{p, 1-p\}.$$

Finally, we get that the expected decrease in expected loss is proportional to

$$V_{jj'}\left[\int_{-\infty}^{D/\rho_b} \phi(x')\Phi\left(\frac{D - \rho_b x'}{\sqrt{1-\rho_b^2}}\right) dx' + \int_{D/\rho_b}^{\infty} \phi(x')\left(1 - \Phi\left(\frac{D - \rho_b x'}{\sqrt{1-\rho_b^2}}\right)\right) dx' - \max\{p, 1-p\}\right]$$

if $\rho_b > 0$ and

$$V_{jj'}\left[\int_{-\infty}^{D/\rho_b} \phi(x')\left(1-\Phi\left(\frac{D-\rho_b x'}{\sqrt{1-\rho_b^2}}\right)\right)dx' + \int_{D/\rho_b}^{\infty} \phi(x')\Phi\left(\frac{D-\rho_b x'}{\sqrt{1-\rho_b^2}}\right)dx' - \max\{p,1-p\}\right]$$

if $\rho_b < 0$, where $\phi$ and $\Phi$ are the the probability density function and cumulative distribution function of the standard-normal distribution. It should be noted that if $p$ is the proportion of positive cases for variable $j$, then $D = \Phi^{-1}(p)$ and that

$$\rho_b = \rho\frac{\sqrt{p(1-p)}}{\phi(D)},$$

where $\rho$ is the correlation between $x$ and $x'$.

##### $x$ and $x'$ are both binary

Let $p$ and $p'$ be the proportion of positive cases in $j$ and $j'$ respectively. Let further $p_{11} = \mathbb{P}(x = 1 \cap x' = 1)$. We know in this case that

$$f_{\text{Bayes}}(x') = \underset{k\in\{0,1\}}{\arg\max}\, \mathbb{P}(x = k|x').$$

This means that it is enough for us to know the contingency table of $x$ and $x'$ to know the Bayes predictor and we know the whole contingency table if we know $p$, $p'$ and $p_{11}$. The maximal expected decrease in the misclassification loss is proportional to

$$V_{jj'}[p'\max\{\mathbb{P}(x=1|x'=1), 1-\mathbb{P}(x=1|x'=1)\} + (1-p')\max\{\mathbb{P}(x=1|x'=0), 1-\mathbb{P}(x=1|x'=0)\} - \max\{p, 1-p\}].$$

The quantity $p_{11}$ is calculated in the following way:

$$p_{11} = \mathbb{E}[xx'] = \text{Cov}[x,x'] + \mathbb{E}[x]\mathbb{E}[x'] = \rho\sqrt{\text{Var}[x]\text{Var}[x']} + pp' = \\ = \rho\sqrt{p(1-p)p'(1-p')} + pp'.$$
